# The predictive value of microfilariae-based stop-MDA thresholds after triple drug therapy with IDA in treatment-naive Indian settings

**DOI:** 10.1101/2023.09.22.23292895

**Authors:** Ananthu James, Luc E. Coffeng, David J. Blok, Jonathan D. King, Sake J. de Vlas, Wilma A. Stolk

## Abstract

Mass drug administration (MDA) of antifilarial drugs is the main strategy towards the elimination of lymphatic filariasis (LF). Recent clinical trials indicated that the triple drug therapy with ivermectin, diethylcarbamazine and albendazole (IDA) is much more effective against LF than the widely used two-drug combinations (albendazole plus either ivermectin or diethylcarbamazine). For IDA-based MDA, the stop-MDA decision is taken based on microfilariae (mf) prevalence in adults. In this study, we assess how the probability of eventually reaching elimination of transmission depends on the critical threshold used in transmission assessment surveys (TAS-es) to define whether transmission was successfully suppressed and triple-drug MDA can be stopped. This analysis focuses on treatment-naive Indian settings. We do this for a range of epidemiological and programmatic contexts, using the established LYMFASIM model for transmission and control of LF. Based on our simulations, a single TAS one year after the last MDA round provides limited predictive value of having achieved suppressed transmission, while a higher MDA coverage increases elimination probability, thus leading to a higher predictive value. Every additional TAS, conditional on previous TAS-es being passed with the same threshold, further improves predictive value for low values of stop-MDA thresholds. An mf prevalence threshold of 0.5% corresponding to TAS-3 results in ≥95% predictive value even when the MDA coverage is relatively low.

## Introduction

Lymphatic filariasis (LF), a neglected tropical disease (NTD), is a leading cause of preventable morbidity and disability due to lymphoedema and hydrocele and still affects over 50 million people worldwide [1]. The most common causative agent is the parasitic filarial nematode worm *Wuchereria bancrofti*. Adult worms are found in lymph vessels, whereas the worm’s offspring microfilariae (mf), released by fertilised female worms, are picked up from blood and transmitted to other humans by mosquitoes. The Global Programme to Eliminate Lymphatic Filariasis (GPELF) was initiated in 2000 with the aim to interrupt transmission of LF by implementing mass drug administration (MDA) with the two-drug combinations of diethylcarbamazine and albendazole (DA) or ivermectin and albendazole (IA) in all affected areas [2–3]. This is to be combined with improved morbidity management to alleviate the suffering of people with clinical manifestations.

Recent clinical evidence indicated that MDA with a triple drug regimen of ivermectin, diethylcarbamazine, and albendazole (IDA) is even more effective than the two-drug regimens [4–7], leading to the World Health Organization (WHO) recommendation in 2017 of using IDA for LF control in LF-endemic areas not co-endemic with either onchocerciasis or loiasis [8]. This includes India, where ∼55% of the current global burden of LF is located, with ∼487 million people at the risk of infection [3]. MDA was initiated in 2004 in 202 Indian districts with DEC and in 2007 in all the 256 endemic districts at that time with DA [9]. In 2018, IDA was introduced in five districts that either failed TAS or never undertook MDA despite the identification of LF transmission. This was followed by the selection of 21 additional districts for IDA implementation in the next year. In 2020, 16 districts were newly classified as endemic for LF, increasing the total number to 272.

MDA programs employing the various drug regimens have successfully reduced LF prevalence in many affected areas so that the infection is either eliminated as a public health problem (mf prevalence <1% or antigen prevalence <2%) or is close to achieving it. To save resources and time, it is important to establish threshold prevalences below which MDA can be stopped as soon as possible but with minimal risk of disease resurgence. For areas treated with a two-drug regimen, the stop-MDA decision is made based on a transmission assessment survey (TAS-1) that assesses the prevalence of the circulating filarial antigenaemia (CFA) in 6-7-year-old children [10]. Elimination is considered validated if antigenaemia prevalence in this age group is sustained below the predefined threshold in 2 subsequent transmission assessment surveys (TAS-2 and TAS-3), with intervals of minimum 2 years between different surveys. However, compared to two-drug regimens, IDA leads to a faster mf clearance and potential sterilisation of adult worms, resulting in fewer required treatment rounds. This means that after the last MDA round, antigens are likely to persist [11]. WHO recognizes that new diagnostics are needed and TPP’s have been developed [12–13]. Until such new tests are available, testing for mf is the best way to identify persons with reproducing adult worms. Because adults, in contrast to children, are the most likely to be mf-positive and have the lowest MDA coverage, it might be useful to target TAS in IDA-treated areas at adults [14].

Mathematical models have been helpful in assessing the validity of stop-MDA thresholds [15–16]. Here we employ the already established LYMFASIM model [16] to simulate IDA-MDA in previously treatment-naïve Indian settings, and to assess the probability of achieving elimination of LF transmission in relation to the chosen mf-based stop-MDA threshold used in TAS-1, TAS-2 and TAS-3. To understand the impact of the employed treatment regimen, including the uncertainty regarding the effect of IDA on adult worms [17], we compare the results for MDA with IDA and DA. We identify situations for which predictive values ≥95% are possible.

## Methods

We adopted the same methods as used in a previous publication on assessing stop-MDA thresholds for the African context [16]. To simulate population-level LF transmission dynamics, we used the individual-based LYMFASIM model, which accounts for inter-host variation in exposure to transmission and uptake of MDA. The model was quantified for Indian settings using data from ∼25,000 individuals from Pondicherry in 1981 and longitudinal measurements of human infection status in 1981, 1986, 1989, and 1991 [18]. This Indian model variant includes a host immunity mechanism that regulates parasite establishment [18–19], which is absent for the African version of LYMFASIM [16]. As a result, transmission can better maintain itself at low levels without incoming infections from neighbouring areas. For the analysis presented here, we simulated a single community with a population of ∼1000 people.

We performed simulations for a range of baseline mf prevalences between 5% and 20%, assuming that simulated communities underwent no previous treatment. Baseline mf prevalences were determined by two transmission parameters: the monthly biting rate and shape of the gamma distribution describing exposure variation between individuals. We repeatedly sampled values for these two parameters from a predefined parameter space (Fig S1); individual sets of parameter values were adopted until we had 250 parameter combinations for each 1%-wide bin between 5% and 20% (i.e., 15 bins). Next, we simulated the impact of 3 to 6 rounds of annual MDA, using either IDA or DA, implemented at either 65% or 80% of the total population, comprising all the age groups (i.e., 16 MDA scenarios). We assumed the macrofilaricidal effect of IDA to be the same as that of DA (55%), but with 100% mf killing and permanent sterilisation of female adult worms (denoted as “IDA1” by Irvine et al. [17]). As such, we ran a total of 3,750 simulations per MDA scenario (15 x 250) and 60,000 simulations in total (16 x 3,750). Each simulation was marked as having achieved elimination (or not) if 20 years after the last MDA round, the mf prevalence in the entire population was zero (above zero). To evaluate the predictive value of TAS surveys for achievement of elimination, for each simulation we saved the annual post-MDA mf prevalence, starting 1 year after the last MDA round. We also repeated TAS-1 defining elimination within a 50-year period post-MDA instead of 20.

We evaluated the predictive value of a series of three TAS-es that take place one, three, and five years after the last MDA round, which was first quantified in terms of receiver operating characteristic (ROC) curves. For each LYMFASIM simulation (N=60,000), we simulated 100 repeated series of TAS-es, assuming a binomial sample of 200 or 400 individuals of ages ≥5 or ≥15 (i.e., 4 TAS scenarios). For each simulated TAS scenario, varying the stop-MDA threshold, we evaluated the positive predictive values (PPV; i.e., the probability of achieving elimination if the measured prevalence is below the threshold), where a particular threshold value was always used for all TAS-es in the same series. (We did not evaluate negative predictive values since overtreatment was outside the scope of this study.) For TAS-2 and TAS-3, we evaluated the PPV on the condition that previous TAS-es in the same series had been passed. We quantified the impact of TAS(-es) in terms of the maximum increment in PPV as the decision threshold is lowered. Finally, we determined the maximum stop-MDA threshold that achieves a PPV of ≥95%. We repeated this analysis with one of the TAS-es being delayed by two years.

## Results

First, we evaluated the amount of information available from TAS-1 targeting age group 5+ with IDA, using ROC curve (Fig. 1). The curve was fairly close to the diagonal, suggesting TAS-1 to provide only limited information relative to the *a priori* probability of achieving elimination. Even for a wide range of MDA scenarios, the ROC curves showed similar trends (Fig. S2). However, the ROC curve for age group 5+ was always further away from the diagonal than age group 15+, indicating that the former is a better option to measure mf prevalence.

**Figure 1.**
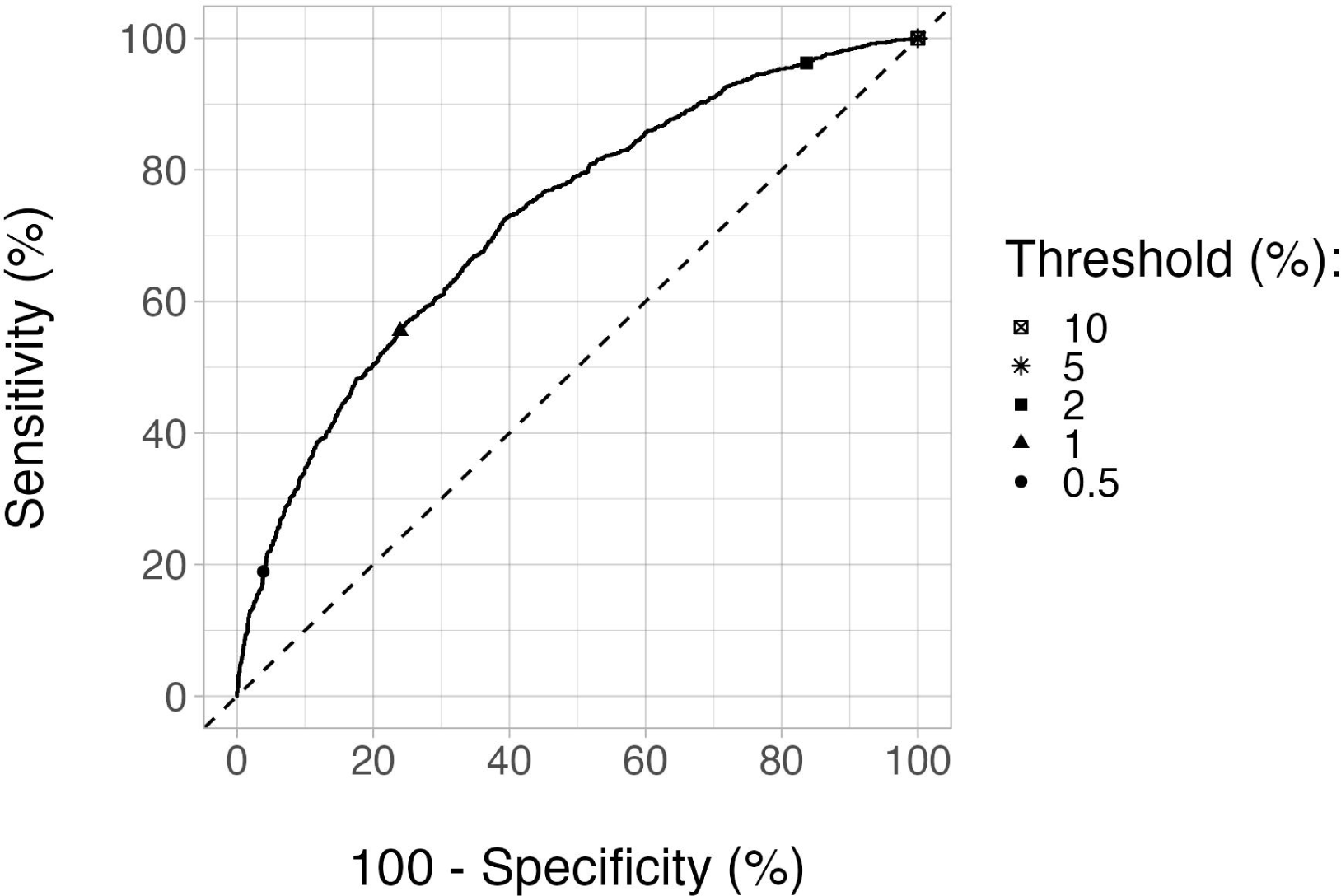
Receiver-operator characteristic (ROC) curve at 1 year post-MDA (TAS-1) for age group 5+ under MDA with IDA. The MDA duration was 3 years and coverage was 65%. The ROC curves for a wide range of parameters can be found in Fig. S2. Note that sampling is not included for the ROC curve(s). (Sensitivity (y-axis) is the percentage of runs ending in elimination within 20 years post-MDA that are correctly identified based on mf prevalence below a range of thresholds. 100%-specificity (x-axis) is the percentage of runs falsely classified as having elimination, among all the runs that did not result in elimination in the same period.)

Next, we evaluated how the PPV of TAS-1 changes as a function of the stop-MDA threshold for mf prevalence in age group 5+ for IDA-treated areas (Fig. 2). One common feature of the PPV curves was that as the stop-MDA threshold increased, the PPV decreased to a plateau, equaling the *a priori* probability of achieving elimination across all simulation runs. For IDA-based MDA scenarios, adopting a lower stop-MDA threshold increased the PPV (relative to the plateau) up to ∼30 percentage-points (Fig. S3). The corresponding increment associated with DA was ∼48 percentage-points. Also, sampling 400 instead of 200 individuals increased the maximum PPV by at most ∼10 percentage-points for IDA (Fig. 2). As the probability of elimination increased with MDA coverage and duration, so did the PPV, and to a degree that was comparable to the magnitude of increase in PPV when adopting lower stop-MDA thresholds. Those aged ≥5 had a higher PPV by at most 5 percentage points than those aged ≥15 for IDA (Fig. S3). With 80% coverage, PPVs of 85-95% were possible when the stop-MDA threshold was ≤0.5% mf prevalence, although surveys to assess mf prevalence led to only a small increment in PPV, compared to the *a priori* probability of elimination.

**Figure 2.**
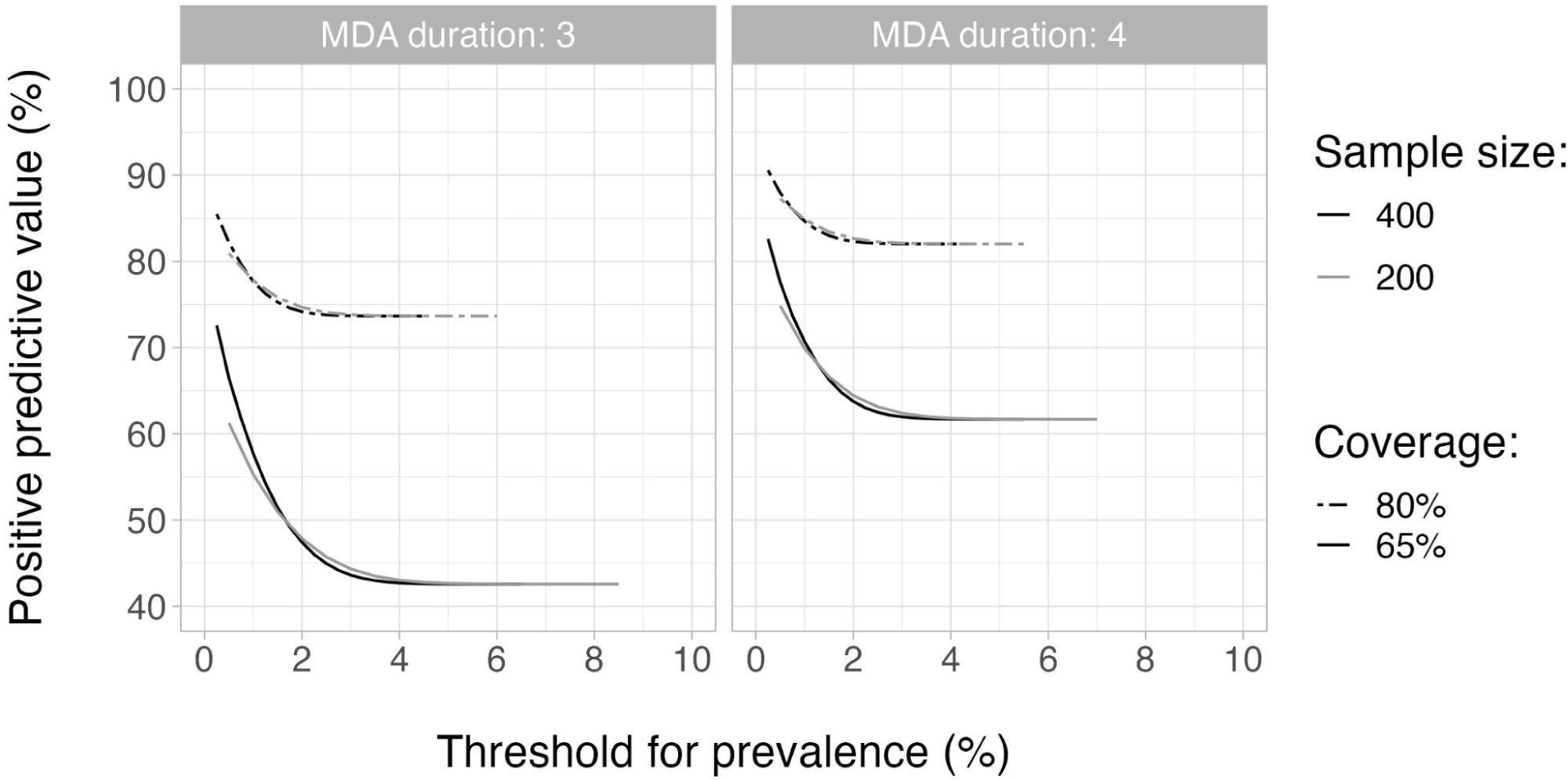
Probability of achieving elimination within 20 years after the last round of IDA-MDA, if the mf prevalence 1-year post MDA (TAS-1) was below a given threshold (i.e., the positive predictive value of using this threshold) for different values of stop-MDA thresholds, for those aged 5+ and for 2 different values of sample sizes (black curve representing 400 and grey curve representing 200), MDA coverages (solid line representing 65% and discontinuous line showing 80%) and durations (3 and 4 years, shown in the left and right panels). The PPV curves, with sample size being 400, for a wide range of other parameters can be found in Fig. S3.

For TAS-1, we considered some alternate situations as well. First, given that the exact values of coverage are often unknown in actual settings, we repeated the above analysis by lumping together simulations with 65% and 80% coverage levels (Fig. S4). The resulting PPV increments were intermediate (with the maximum increments being ∼25 percentage-points for IDA and ∼42 percentage-points for DA) between those when the two coverage levels were considered separately. Next, to know if the predictive power of TAS-1 would improve, we considered elimination within 50 years instead of 20 years (Fig. S5). The *a priori* probability of elimination was higher if measured 50 years after stopping than if measured 20 years after stopping, resulting in higher absolute PPVs. However, the maximum PPV increments were lower for the former for both the drug regimens, indicating its weaker predictive power. The maximum PPV increments were ∼20 and ∼30 percentage points for the two periods for IDA-based MDA, whereas they were ∼35 and ∼48 percentage points for the DA-based one.

Finally, keeping the time to achieve elimination as 20 years, we explored the predictive value of TAS-2 and TAS-3, conditional on previous TAS-es being passed (with the same stop-MDA threshold as used in later TAS-es), when based on a sample of ages 5+ (Fig. 3). PPV increased with each additional TAS for lower thresholds. This trend remained consistent across all the MDA scenarios we considered (Fig. S6). Following TAS-2 and TAS-3, the maximum PPV increments associated with IDA became ∼45 and ∼52 percentage points, respectively, compared to the maximum increase of ∼30 percentage points for TAS-1 only. The corresponding values for DA were ∼68 and ∼76 percentage points, respectively, compared to the ∼48 percentage points for TAS-1 only. Thus, PPVs following TAS-3 can be as high as ∼95% for decision thresholds ≤0.5% mf prevalence, even for low values of MDA coverage (65%) and duration (three years).

**Figure 3.**
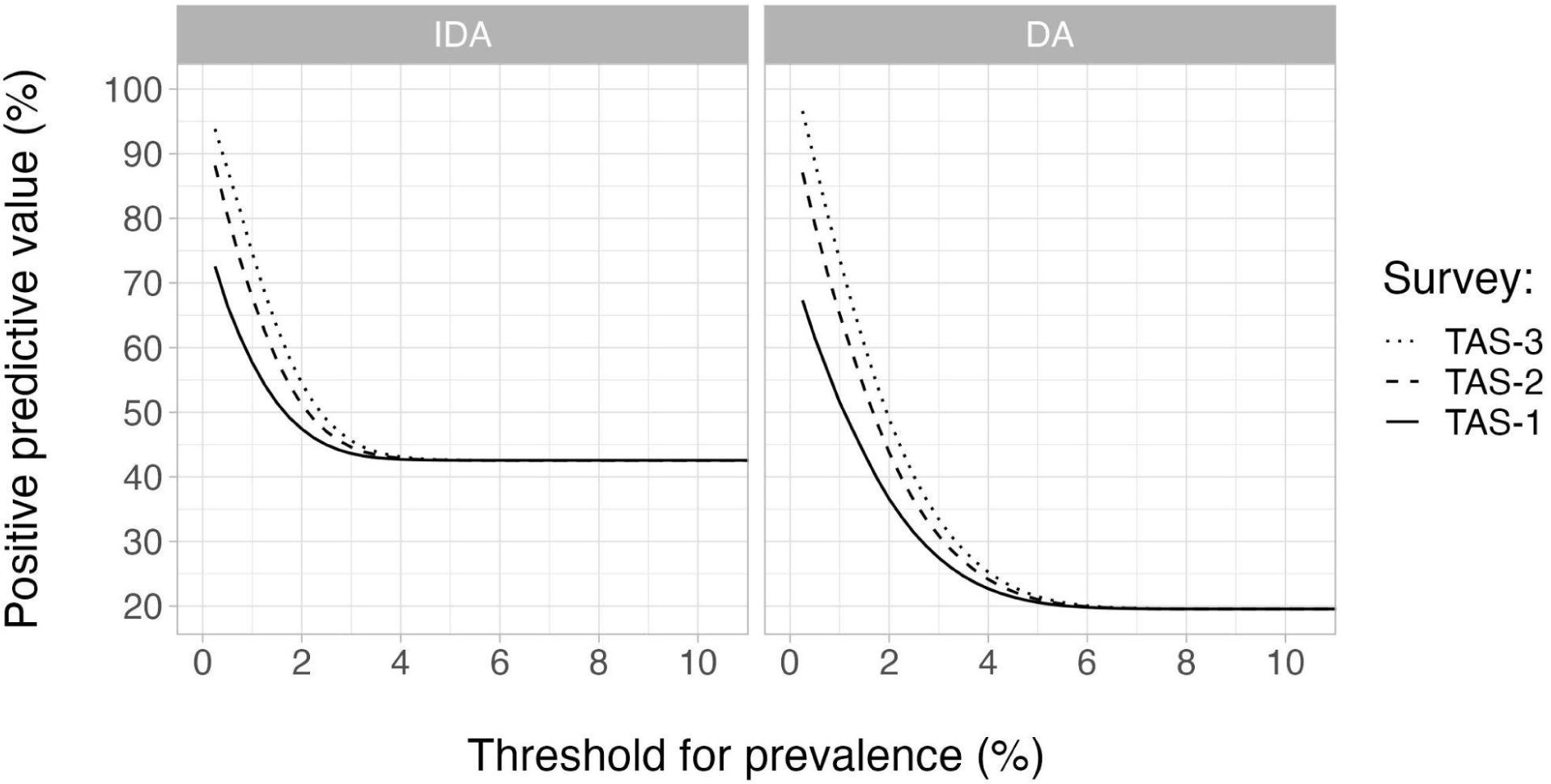
Positive predictive value (PPV) of TAS-1, TAS-2, and TAS-3 for elimination of LF after 3 years of MDA at 65% coverage. Elimination was defined as zero mf prevalence 20 years after the last MDA round. PPVs were calculated as a function of the stop-MDA threshold for prevalence of infection in the age group 5+ (horizontal axis). For TAS-2 and TAS-3, PPVs are conditional on all previous TAS-es being passed with the same prevalence threshold. TAS-1, 2, and 3 were scheduled 1, 3, and 5 years post-MDA, so that the gap between consecutive TAS-es was 2 years. The left panel represents MDA with IDA and the right one represents DA. For each TAS, we assumed a sample size of 400.

We also found that delaying TAS-1 by two years, so that TAS-es occurred at three, five, and seven years post-MDA, had only a limited impact on PPVs (Fig. S7). The PPV increment was the highest for TAS-1, by ∼5 percentage points, for both the drug regimens. The PPV increments associated with increasing the gap between one of the TAS-es, keeping TAS-1 at 1-year post-MDA, were even more negligible (not shown).

## Discussion

Our results show that by itself, TAS-1 conducted to measure mf prevalence 1-year after the last MDA round with IDA provides relatively little information on the prospect of elimination, which is dictated largely by MDA duration and coverage. However, TAS-2 and TAS-3, conducted 3 and 5 years post-MDA (but only if TAS-1 is passed), are more informative in this regard. For TAS-3, PPVs ≥95% are possible with a stop-MDA threshold of ≤0.5% mf prevalence in ages 5 and above. Further, using a larger sample size for TAS (i.e., more number of individuals) as well as testing the age group of 5+ instead of 15+ increase the PPV of mf surveys. The latter is due to the fact that if MDA successfully suppresses transmission, young individuals are less likely to contract their first worm infection; therefore, in age groups where prevalence would normally strongly increase with age pre-MDA (ages 5-15), a low infection prevalence is indicative of a significant impact on transmission. Including this age group in surveys therefore adds useful information for decision making [15].

The surveys provide more information in settings with DA than setting with IDA, which is due to IDA being more effective in clearing LF infection than DA. Results pertaining to DA are helpful in hypothesising the consequence of an alternate assumption of IDA having no sterilising effect (denoted as “IDA2” by Irvine et al. [17]), contrary to the 100% (permanent) sterilisation of female adult worms we considered here. In case of no sterilising effect, the only difference of IDA2 from DA is its ability of killing 100% mf instead of the widely accepted value of 95% mf killing of DA [20]. Therefore, we expect that the results for an analysis of IDA2 would be similar to what we presented here for DA.

Previous modelling studies on elimination or stop-MDA thresholds for LF considered 40-50 years after the last MDA round as the time horizon to define elimination (0% mf prevalence) [16, 19, 21], whereas we adopted a time horizon of 20 years. Following the assumption that the parasite numbers are so low by the time MDA is stopped, as per the breakpoint theory [22], the number of new infections post-MDA is not enough to sustain transmission. Hence, when past this breakpoint, a 50-year period would allow for the complete natural attrition of the remaining parasite population in our simulations, resulting in a higher (*a priori*) elimination probability than for a 20-year period. This is why TAS-es added less information in the former case than the latter. More importantly though, a 50-year time horizon is far beyond the political scope of most governments and probably not realistic anyway since a lot can happen in 50 years (e.g., secular developments or disaster). Although still long, a 20-year time horizon is much closer to the reality in which we develop and employ NTD control strategies. For this time horizon, it is rather more feasible to also compare our model predictions with results from actual settings than for the 50-year period.

In real-life settings, before TAS-1, a pre-TAS is often conducted in up to eight communities to assess whether mf prevalence is <1% (CFA prevalence <2%). The purpose of the pre-TAS is to decide whether TAS-1 should be conducted across the evaluation unit (typically a district), which would encompass a large number of communities. As our simulations focus on single communities and not larger areas consisting of multiple communities, we did not account for pre-TAS. Also, we did not account for the fact that mf tests are likely not performed population-wide due to the inconvenient need to use night blood samples. In reality, programmes will likely perform antigen surveys during the day, with mf tests only carried out in antigen-positives, as in recent studies like the one by Eneanya *et al.* [23]. In our study, we assume that the probability of an mf-positive individual testing negative on CFA is negligible, and that as such, our results can be taken to represent the aforementioned testing strategy, that is, if decision-making is based solely on the estimated mf prevalence. If the CFA prevalence itself would also be considered in the decision-making (e.g., via a second threshold for CFA prevalence), the predictive value of TAS-es might be even somewhat higher than we predict here. Further, the timing of TAS-es in reality may not be as we assumed here (every two years, starting 1 year after the last MDA round). For instance, it is also possible that TAS-es could be delayed in some settings due to external reasons such as COVID-19. However, we found such delays to only slightly increase the predictive value of TAS-es, which is reflective of the slow dynamics of LF recrudescence [24].

To assess the status of LF elimination, only mf and CFA prevalences are used as indicators, with the latter not being recommended for IDA-based MDA. For other NTDs, vector infectivity rate is also often used as an indicator. For example, stop-MDA decisions for onchocerciasis are based on black fly infectivity in addition to antibody seroprevalence [25]. However, in the case of LF, there is no such established practice of collecting mosquito infectivity data. This is also because the mosquitoes would need to be captured in many different locations (e.g., households) to obtain a representative picture of a community [26], unlike the onchocerciasis transmitting black flies which circulate throughout the village making it easy for their data to be collected.

In this work, we only included regions in India that were previously untreated. Still, there might well be settings that failed TAS even after multiple rounds of DA and where IDA could be administered for 1-3 rounds, for which a similar analysis would be interesting. IDA can also be administered in LF endemic African regions that are not endemic to both onchocerciasis and loasis, such as Madagascar. Our analysis could be extended to such settings as well.

We conclude that when only TAS-1 is included, PPVs are always <95% for threshold values ≥0.5% mf prevalence. However, with two additional TAS-es, spaced by two years and conditional on all three TAS-es being passed with the same threshold, PPVs of ≥95% are possible for a stop-MDA threshold of 0.5%, even when the coverage is as low as 65%. This study supports the WHO strategy of repeating the TAS twice during post-MDA surveillance, although the PPV could be improved by lowering the decision threshold from ∼4% to 0.5%.

## Data Availability

The data relevant for this work were quantified and described in detail in previous publications. These articles were mentioned in the Methods section of the article.

## Acknowledgements

This work was supported by funding of the NTD Modelling Consortium by the Bill & Melinda Gates Foundation. This supplement is sponsored by funding of Professor T. Déirdre Hollingsworth’s research by the Li Ka Shing Foundation at the Big Data Institute, Li Ka Shing Centre for Health Information and Discovery, University of Oxford and funding of the NTD Modelling Consortium by the Bill & Melinda Gates Foundation (INV-030046). The funders had no role in study design, data collection and analysis, decision to publish, or preparation of the manuscript.

---

**Figure S1.**
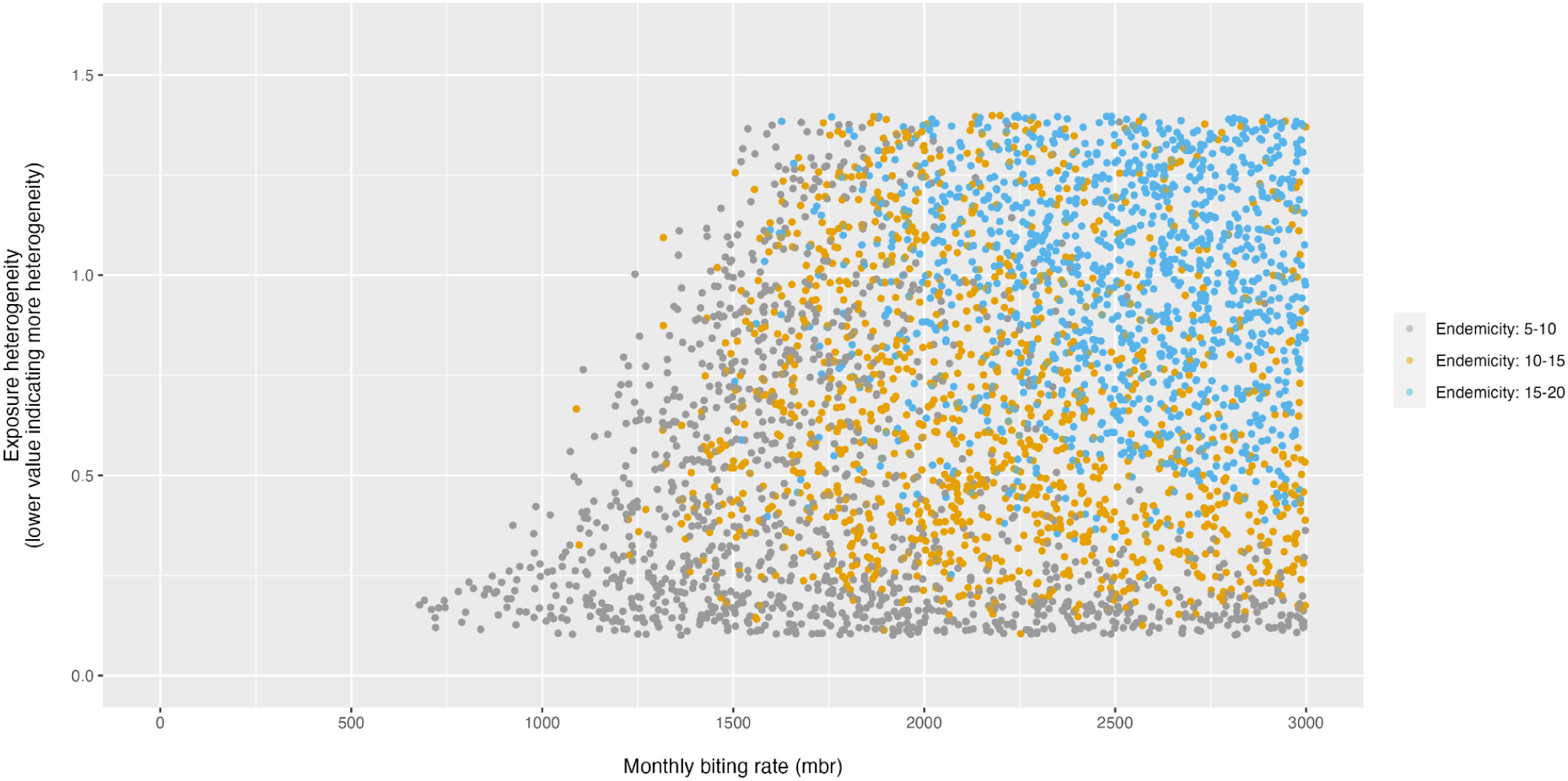
Parameter combinations used for simulations, coloured by baseline endemicity class, with endemicity defined in terms of mf prevalence. Note that these endemicity classes do not appear in the results, since all the simulations in this article are performed after lumping endemicity classes together.

**Figure S2.**
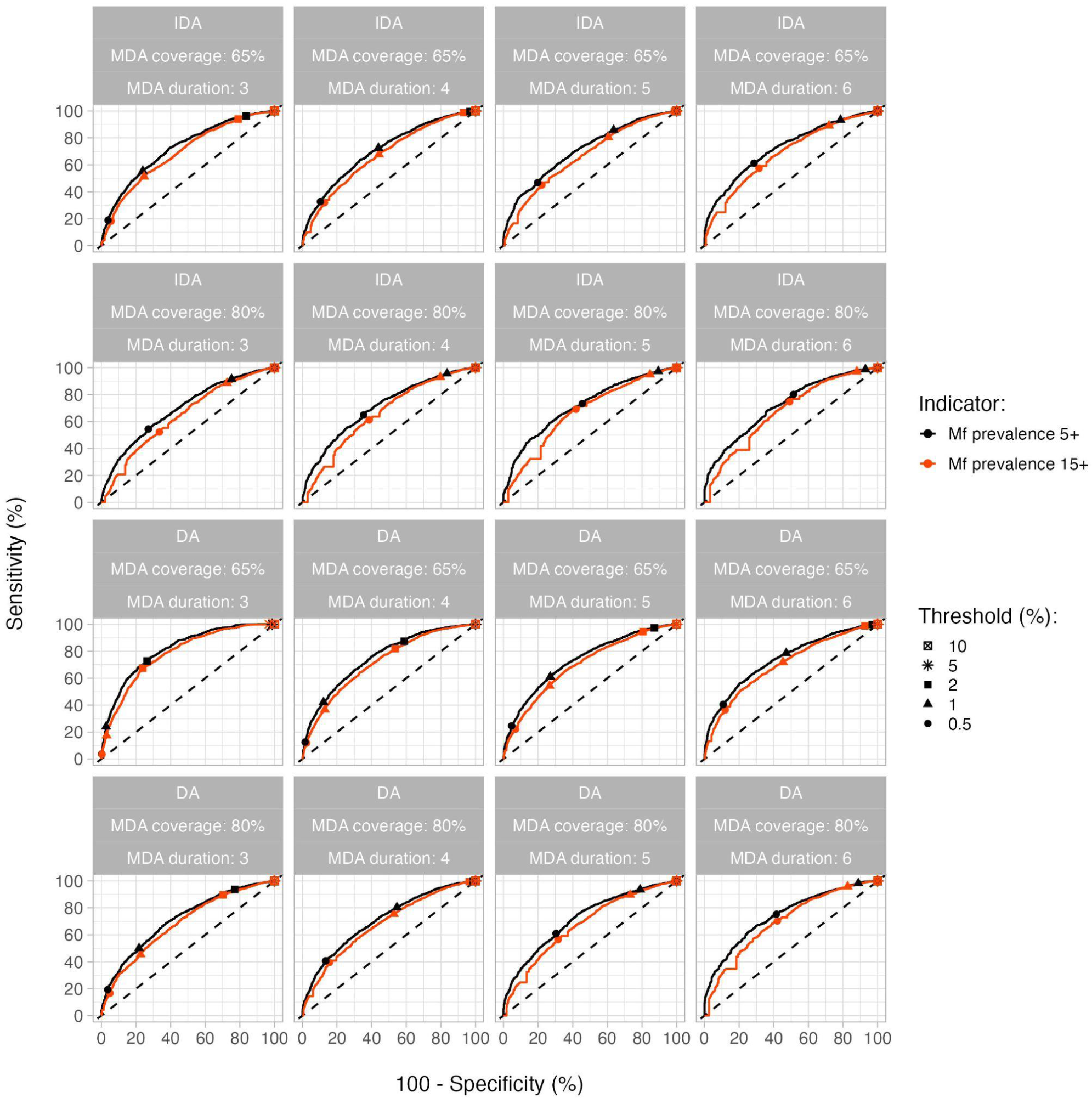
Receiver-operator characteristic (ROC) curves for two different age groups 1 year post-MDA (TAS-1), when elimination was defined within 20 years post-MDA. The panels in the top two rows represent MDA with IDA and the bottom ones MDA with DA.

**Figure S3.**
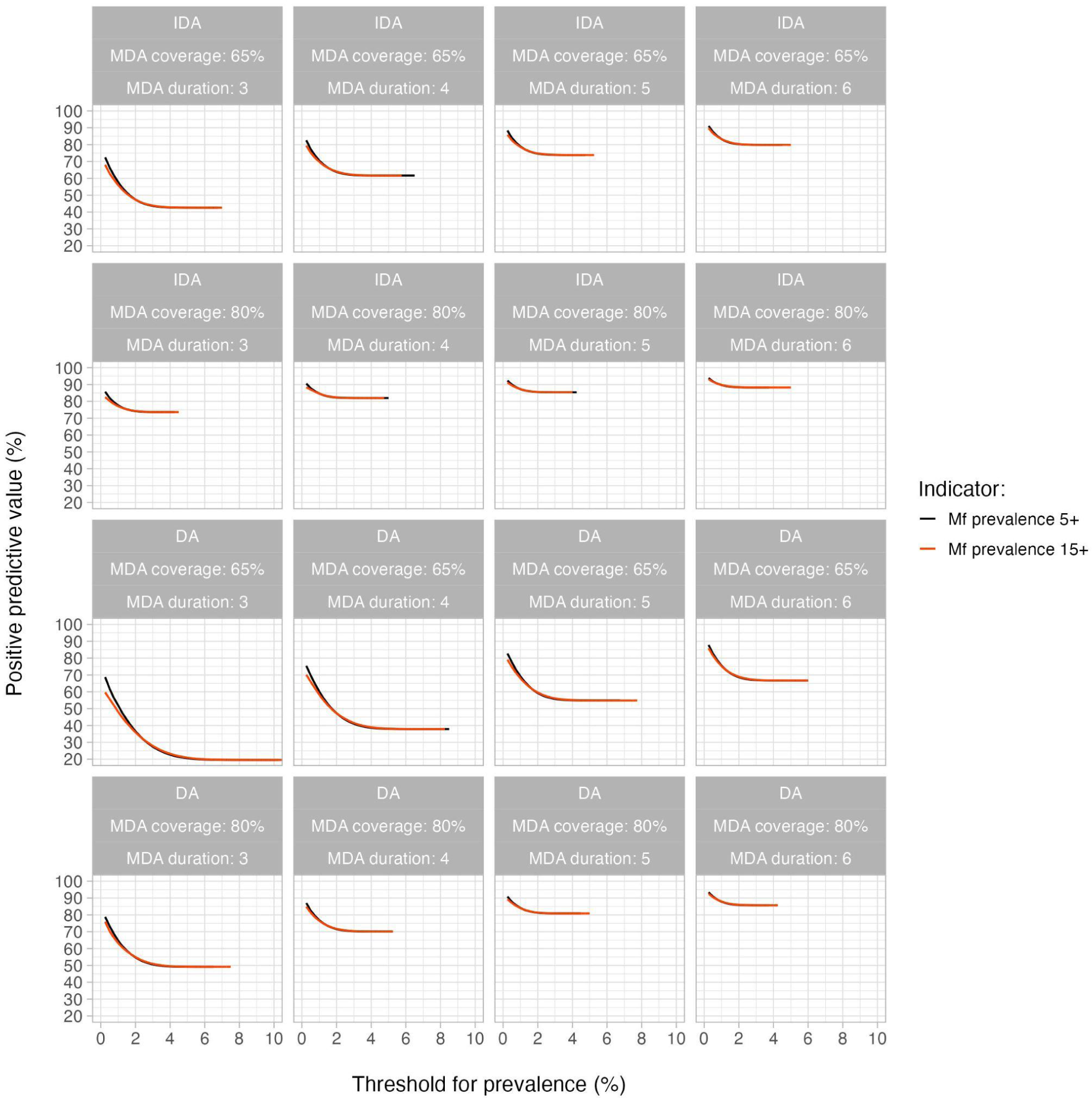
Positive predictive value (PPV) of TAS-1 for elimination of LF for different values of MDA duration and coverage. Elimination was defined as zero mf prevalence 20 years after the last MDA round. PPVs were calculated as a function of the stop-MDA threshold for prevalence of infection (horizontal axis) in the age groups 5+ (black curve) and 15+ (red curve). The panels in the top two rows represent MDA with IDA and the bottom two rows represent MDA with DA. We assumed a sample size of 400.

**Figure S4.**
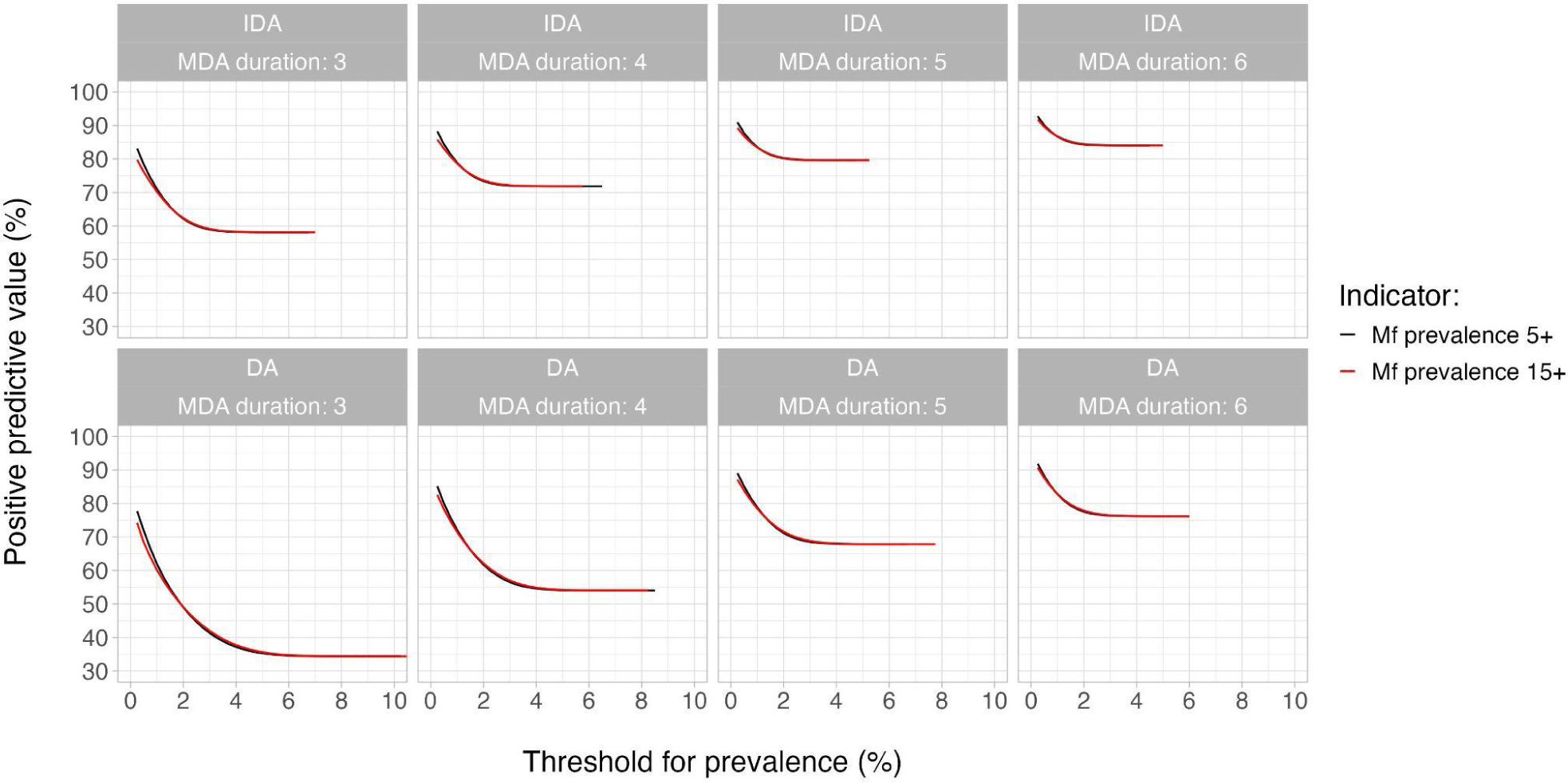
Positive predictive value (PPV) of TAS-1 for elimination of LF for different values of MDA duration when coverages were lumped together for different age groups. Elimination was defined within 20 years post-MDA. PPVs were calculated as a function of the stop-MDA threshold for prevalence of infection (horizontal axis) in the age groups 5+ (black curve) and 15+ (red curve). The top panels represent MDA with IDA and the bottom panels represent DA. The sample size was 400.

**Figure S5.**
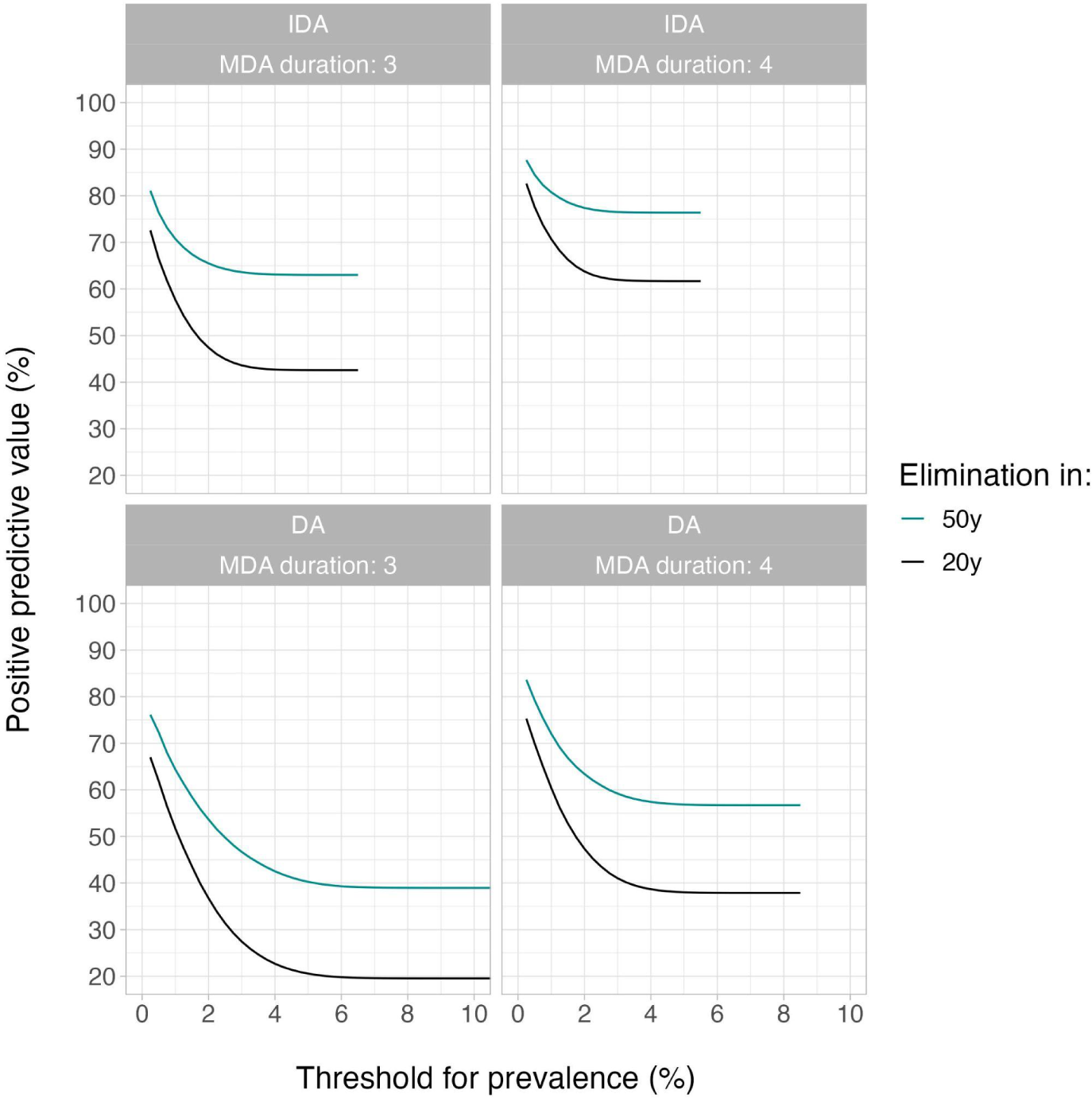
Positive predictive value (PPV) of TAS-1 for elimination of LF for 65% MDA coverage and two different values of duration. Elimination was defined within 20 years (black curve) and 50 years (cyan curve) post-MDA. PPVs were calculated as a function of the stop-MDA threshold for prevalence of infection (horizontal axis) in the age groups 5+. The top panels represent MDA with IDA and the bottom ones represent DA. The sample size was 400.

**Figure S6.**
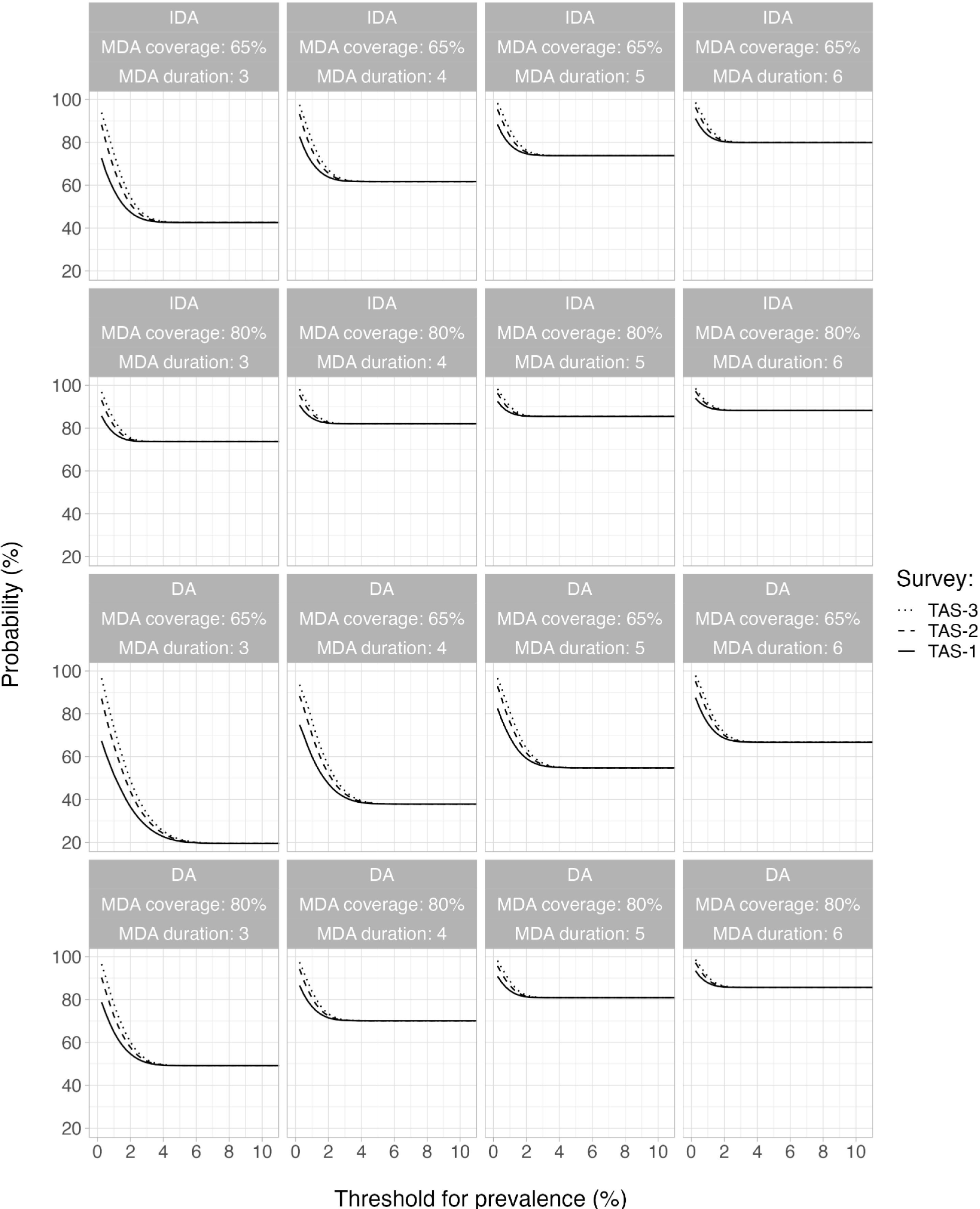
Positive predictive value (PPV) of TAS-1, TAS-2, and TAS-3 for elimination of LF for different values of MDA duration and coverage. Elimination was defined as zero mf prevalence 20 years after the last MDA round. PPVs were calculated as a function of the stop-MDA threshold for prevalence of infection in the age group 5+ (horizontal axis). For TAS-2 and TAS-3, PPVs are conditional on all previous TAS-es being passed with the same prevalence threshold. TAS-1, 2, and 3 were scheduled 1, 3, and 5 years post-MDA, so that the gap between consecutive TAS-es was 2 years. The panels in the top two rows represent MDA with IDA and the bottom two represent DA. For each TAS, we assumed a sample size of 400.

**Figure S7.**
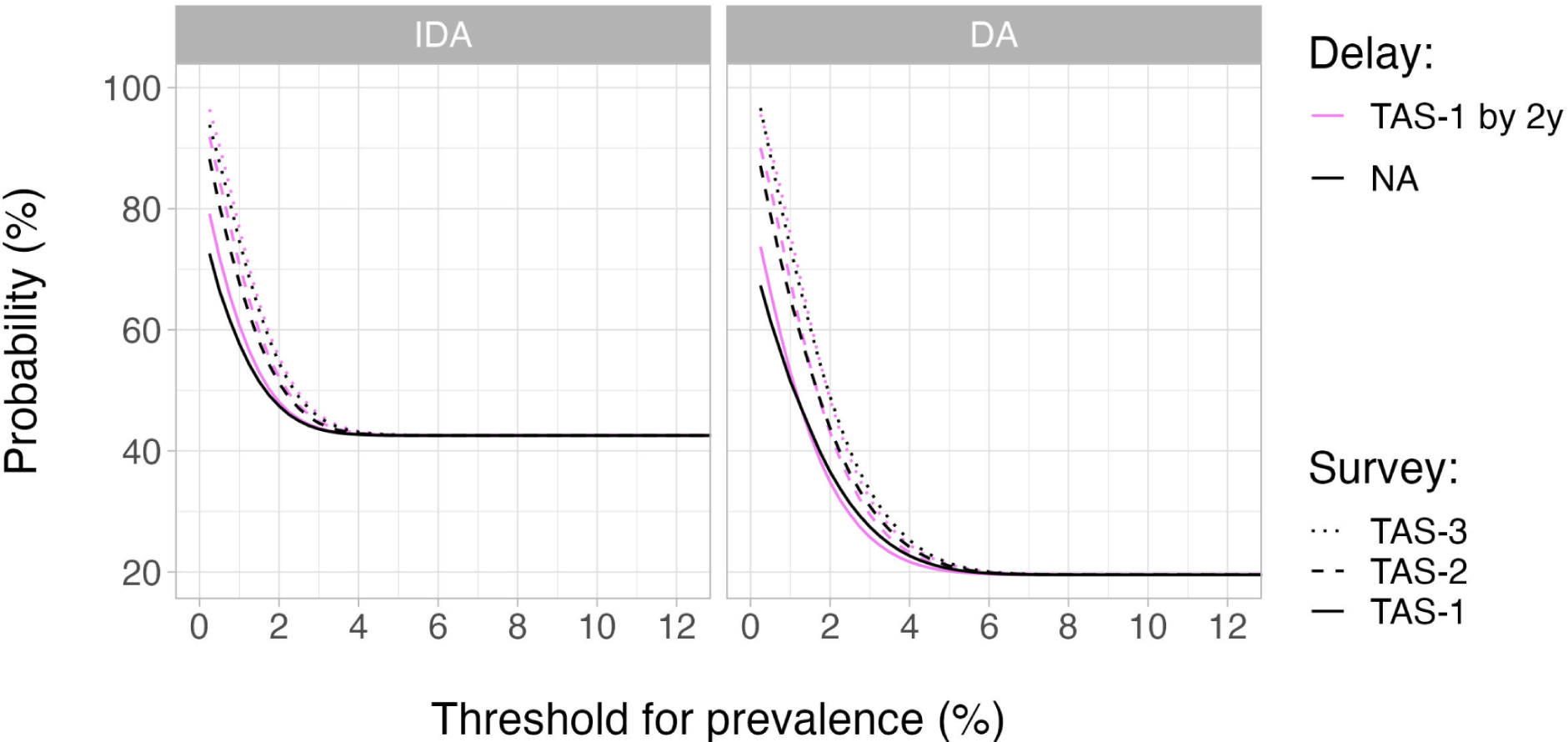
Positive predictive value (PPV) of TAS-1, TAS-2, and TAS-3 for elimination of LF following MDA for 3 years at 65% coverage. Elimination was defined as zero mf prevalence 20 years after the last MDA round. PPVs were calculated as a function of the stop-MDA threshold for prevalence of infection in the age group 5+ (horizontal axis). For TAS-2 and TAS-3, PPVs are conditional on all previous TAS-es being passed with the same prevalence threshold. We considered two situations for the surveys: (i) TAS-1, 2, and 3 being scheduled 1, 3, and 5 years post-MDA (black curves) and (ii) TAS-1, 2, and 3 being scheduled 3, 5, and 7 years post-MDA (violet curves). The left panel represents MDA with IDA and the right one represents DA. For each TAS, we assumed a sample size of 400.

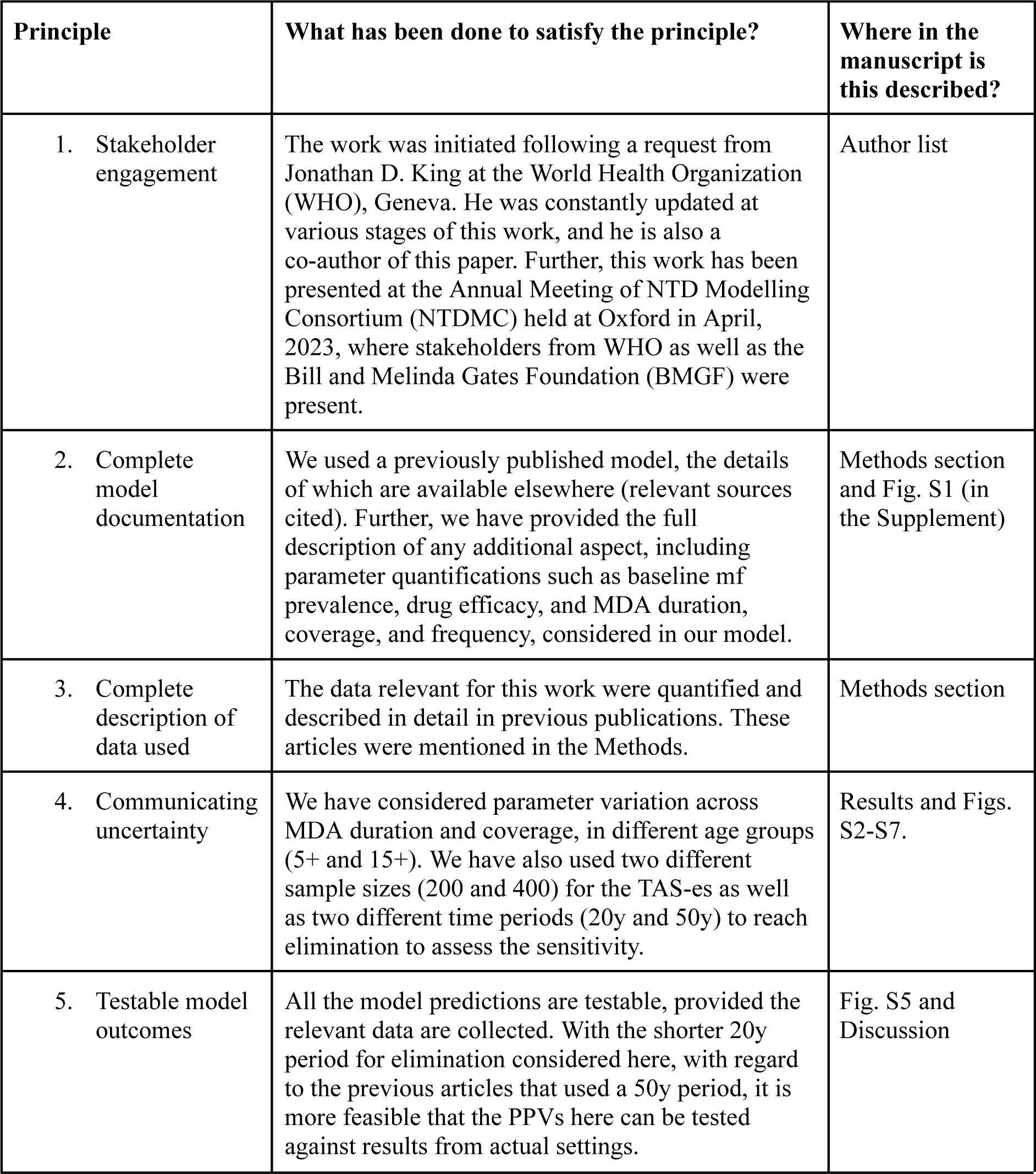
PRIME-NTD table: Policy-Relevant Items for Reporting Models in Epidemiology of Neglected Tropical Diseases.

| Principle | What has been done to satisfy the principle? | Where in the manuscript is this described? |
| --- | --- | --- |
| 1. Stakeholder engagement | The work was initiated following a request from Jonathan D. King at the World Health Organization (WHO), Geneva. He was constantly updated at various stages of this work, and he is also a co-author of this paper. Further, this work has been presented at the Annual Meeting of NTD Modelling Consortium (NTDMC) held at Oxford in April, 2023, where stakeholders from WHO as well as the Bill and Melinda Gates Foundation (BMGF) were present. | Author list |
| 2. Complete model documentation | We used a previously published model, the details of which are available elsewhere (relevant sources cited). Further, we have provided the full description of any additional aspect, including parameter quantifications such as baseline mf prevalence, drug efficacy, and MDA duration, coverage, and frequency, considered in our model. | Methods section and Fig. S1 (in the Supplement) |
| 3. Complete description of data used | The data relevant for this work were quantified and described in detail in previous publications. These articles were mentioned in the Methods. | Methods section |
| 4. Communicating uncertainty | We have considered parameter variation across MDA duration and coverage, in different age groups (5+ and 15+). We have also used two different sample sizes (200 and 400) for the TAS-es as well as two different time periods (20y and 50y) to reach elimination to assess the sensitivity. | Results and Figs. S2-S7. |
| 5. Testable model outcomes | All the model predictions are testable, provided the relevant data are collected. With the shorter 20y period for elimination considered here, with regard to the previous articles that used a 50y period, it is more feasible that the PPVs here can be tested against results from actual settings. | Fig. S5 and Discussion |

